# A mixed methods evaluation of 99DOTS digital adherence technology uptake among adolescents treated for pulmonary tuberculosis in Uganda

**DOI:** 10.1101/2024.12.01.24318270

**Authors:** P Wambi, N.S. West, J Nabugoomu, A Kityamuwesi, R Crowder, L Kunihira, E Wobudeya, A Cattamanchi, D Jaganath, A Katamba

## Abstract

**Introduction:** Adolescents and young adults are at risk of poor adherence to tuberculosis (TB) treatment and subsequently worse TB treatment outcomes. Digital adherence technologies, including the mobile phone-based 99DOTS platform, can support TB treatment, but there is limited data on their use among adolescents.

**Objective:** To evaluate factors associated with the uptake of 99DOTS among adolescents with TB.

**Methods:** We conducted an explanatory sequential mixed methods study that utilized quantitative data from adolescents collected during the scale-up of 99DOTS at 30 health facilities in Uganda, and qualitative in-depth and key informant interviews with a subset of adolescents with pulmonary TB offered 99DOTS and healthcare providers at participating facilities. Findings were further mapped onto the Capability, Opportunity, Motivation, and Behavior (COM-B) model.

**Results:** Overall, 299/410 (73%) eligible adolescents were enrolled in 99DOTS. Older adolescents 15 to 19 years old were more likely to enroll in 99DOTS than younger adolescents 10 to 14 years [aPR= 0.56, 95% CI: (0.42-0.73)]. Conversely, adolescents treated at Health Center IV and General Hospitals were less likely to be enrolled compared to Health Center III (aPR= 0.8, 95% CI, 0.67-0.94, and aPR=0.71, 95% CI 0.58-0.85, respectively). Technological savviness among older adolescents, access to training, caregiver or treatment supporter involvement, and desire for wellness facilitated the uptake of 99DOTS. In contrast, variable mobile phone access, concerns about TB status disclosure, and health worker workload in hospitals were barriers to the uptake of 99DOTS.

**Conclusion:** 99DOTS uptake was high among adolescents with TB. Increased access to mobile phones, health worker training on adolescent communication, and more involvement of caregivers could facilitate greater use of 99DOTS and similar technologies.

## Introduction

Tuberculosis (TB) is the leading infectious disease cause of death worldwide (1). Among 10.6 million people who became ill with TB in 2022, an estimated 1.8 million are adolescents and young adults -ages 10-24 years (1). Adolescents have unique social and biological determinants that increase their risk of TB treatment adherence challenges (2, 3) and subsequent poor treatment outcomes as compared to adults and children (4, 5). However, adolescents are often overlooked as a unique population in TB and are frequently grouped with children or adults (6), limiting the development of adolescent-specific interventions.

Uganda ranks top among high-prevalence TB countries worldwide, with an estimated incidence of 198 per 100,000 population (7). Adolescents and young adults aged 15 to 24 years account for approximately 13% of TB disease in the country, with treatment success rates varying between 50% and 85% depending on the region (7). A study in Uganda found that adolescents with TB were twice as likely to be lost to follow-up compared to children under five years, when receiving standard community-based directly observed therapy (CB-DOT) (5). Further, non-adherence to TB treatment and risk of loss of follow-up are persistently higher among HIV-TB co-infected adolescents (8).

Globally, there is increasing advocacy for investing in and using digital adherence technologies (DATs) within National TB Programs (NTPs) to provide remote treatment support and monitoring for people with TB (Stop TB Partnership 2019). In 2017, the World Health Organization (WHO) recommended the use of information technologies to improve TB treatment outcomes (9). The Uganda National TB Leprosy Program has further prioritized using digital technologies to tackle the challenge of poor treatment adherence and improve treatment outcomes among all patients, including adolescents (10). Recent studies have shown promising results with these technologies for adherence among adults in Uganda (11–13).

99DOTS is a DAT that utilizes low-end mobile phones (i.e. not smart phones) for patients and a web-based dashboard and mobile phone application for healthcare workers. It consists of automated SMS dosing reminders, daily reporting of dose completion by calling a random toll-free telephone number on TB medication blister packs, and weekly automated interactive voice response check-in phone calls. Clinic staff can view real-time adherence data for each patient via the mobile app and web dashboard. In the DOT to DAT stepped-wedged cluster randomized trial in Uganda, we found that 99DOTS was highly feasible and acceptable among adults with pulmonary TB (13–15), and achieved high treatment completion (12).

Beginning in 2021, the 99DOTS intervention was scaled up to 30 health facilities across Uganda, including the health facilities previously enrolled in the DOT to DAT trial, and became available also to adolescents. The objective of this study was to use sequential explanatory mixed methods to evaluate the factors associated with the uptake of the 99DOTS intervention among adolescents on treatment for drug-susceptible pulmonary TB (DS-PTB) treatment in Uganda.

## RESULTS

### PHASE I: QUANTITATIVE

#### Socio-demographic characteristics of study participants

A total of 410 adolescents with TB were included. The median age was 17 years (IQR 14-18), 59% (n=241) were female; 20% (n=82) were living with HIV, 60% (n=246) had microbiologically confirmed TB, and 75.8% had at least one treatment supporter (Table 1).

**Table 1:** Overall socio-demographic and clinical characteristics (n=410)

|  | **Variable** | **Frequency (%)** |
| --- | --- | --- |
| **Age** | 10-14 | 129 (31.5%) |
|  | 15-19 | 281 (68.5%) |
| **Sex** | Female | 241 (58.8%) |
|  | Male | 169 (41.2%) |
| **Disease Classification** | Confirmed TB | 246 (60.0%) |
|  | Clinically Diagnosed TB | 164 (40.0%) |
| **HIV Status** | HIV Negative | 326 (79.5%) |
|  | HIV Positive | 82 (20.5%) |
| **Treatment support** | Yes | 311 (75.8%) |
|  | No | 46 (11.2%) |

#### Uptake of 99Dots

Overall, 299 (73%) were enrolled in 99DOTS during the study period. Of the study participants enrolled on 99DOTS, higher uptake was seen among older adolescents (15-19 years), females, those who had confirmed TB, adolescents who had a caretaker, and those who were enrolled at lower-level health centers and regional referral hospitals (Table 2). The adjusted prevalence of uptake of 99DOTS was higher among older (15-19 years) than younger (10-14 years) adolescents (aPR 1.88 95% CI: 1.54-2.33) (Table 3) and adolescents who had a treatment supporter (*i..e,* family member who was their caretaker) (aPR 1.44, 95% CI: 1.13-1.83). The adjusted prevalence of uptake of 99DOTS was lower among adolescents treated at either Health Center IVs (aPR 0.8, 95% CI: 0.67-0.94) or General Hospitals (aPR = 0.71, 95% CI: 0.58-0.85) compared to adolescents treated at Health Center IIIs (Table 2).

**Table 2:**
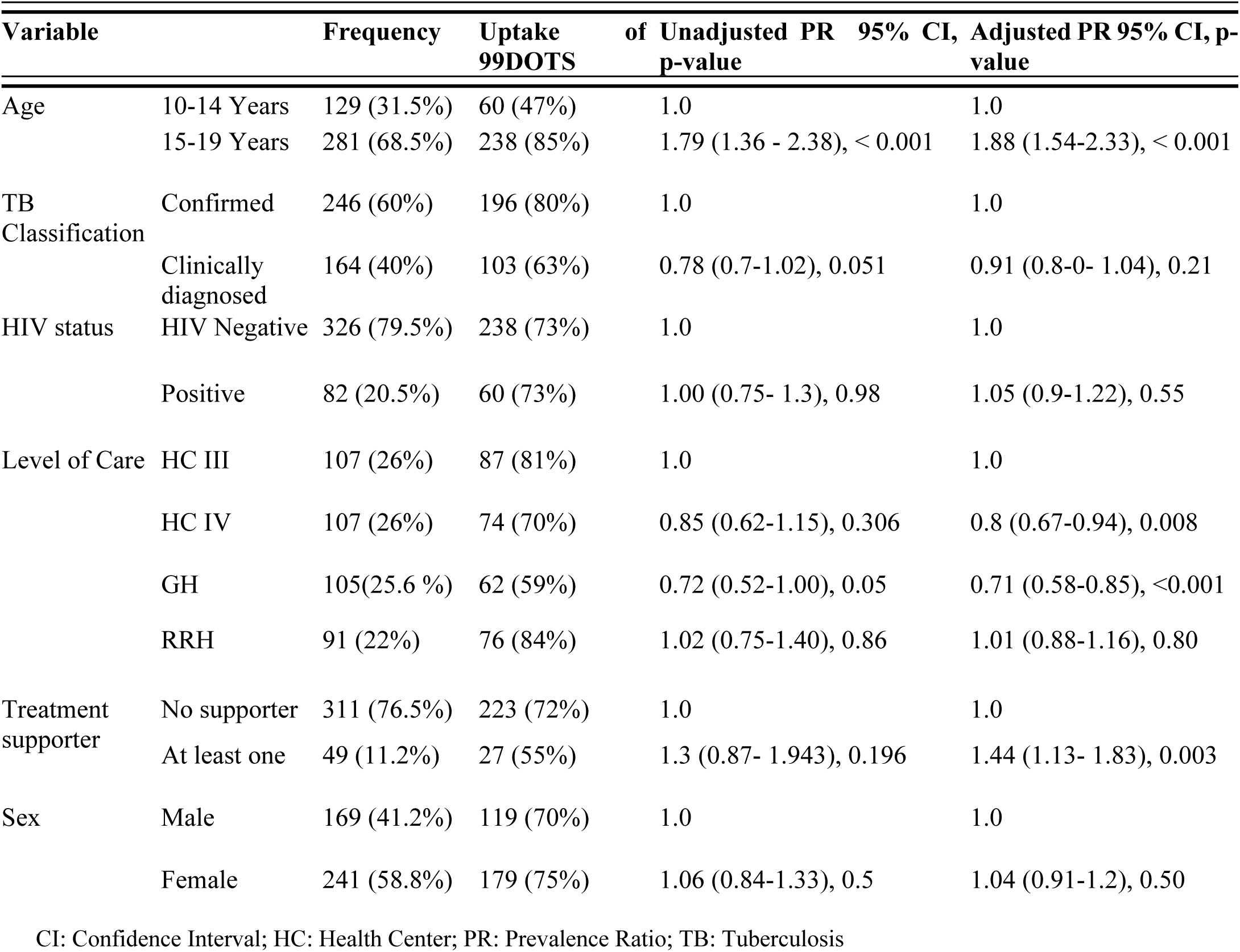
Uptake of 99DOTS stratified by Age, Sex, Disease classification, HIV status, treatment support, and level of care.

### PHASE II: QUALITATIVE

Of the 20 in-depth interviews conducted among a sub-set of adolescents, nine were eligible but did not participate in 99DOTS, and 11 had engaged with 99DOTS. Overall, 50% (n=10) were male, 70% (n=14) were 15-19 years of age, 40% (n=8) were living with HIV, and 45% (n=9) received care at lower-level health facilities (level III or IV). Key informant interviews were conducted with eight health workers. Among the health workers, 50% (n=4) were female, and three were employed at lower-level health facilities. Themes explaining quantitative findings in the context of facilitators and barriers to the uptake of 99DOTS by adolescents were mapped onto each of the three domains of the COM-B model to elucidate the multifaceted factors that influence the uptake of 99DOTS.

#### Facilitators of Uptake of 99DOTS

##### Capability

Existing technological savviness among adolescents, regardless of age, was a psychological facilitator of the uptake of 99DOTS for many adolescents, who described being attracted to the novel mobile technology and were quickly able to grasp how to use 99DOTS. Similarly, technological comfort also enhanced health worker capability to use 99DOTS. Overall, most participants described that adolescents were comfortable with phones and the 99DOTS technology:

> *“The digital world, you know adolescents and the digital world…they were already interested in phones, and therefore it was easy for them, the adolescents were the best people [for the 99DOTS approach]* -Healthcare provider, HC III health facility

##### Opportunity

Caregivers played a key role in supporting opportunities for uptake of 99DOTS by providing phone access and, in some cases, a commitment to support the adolescents in using the system. In addition, many adolescents highlighted that access to clear training and education on the use of 99DOTS provided by healthcare providers facilitated the opportunity to participate in the approach.

##### Motivation

The desire to recover from tuberculosis was an essential driver of uptake of the 99DOTS among adolescents, irrespective of age and HIV status, as described by an adolescent who participated in 99DOTS.

> *“It [99DOTS] gave me the encouragement that I have to take medicine as my life is my life and not anyone else’s; that is it, and I remember some words in Luganda: ‘For your life to be good’*-Adolescent with TB, participated in 99DOTS, 15-19 years old

Most adolescents and healthcare providers said that the 99DOTS novel packaging and design appealed to adolescents and remotely reminded and encouraged them to take their medication. Adolescent participants, in particular, felt the medication packaging was appropriately nondescript and allayed fears about inadvertent disclosure and TB-related stigma. Further, healthcare providers at all levels of care reported that 99DOTS improved their communication with adolescent TB patients in a way that enhanced their ability to care for and communicate with adolescents with TB.

> *“You know adolescents are very stubborn. First of all, they are faced with stigma, and secondly, they are faced with peer issues. They are also faced with the challenge of taking treatment in hiding. When 99DOTS is there, you can monitor them and get closer to them as they are taking treatment.”* -Healthcare provider, GH health facility

Some adolescents also reported that they were motivated to use 99DOTS because of the enhanced or better-facilitated access to their healthcare providers that the approach offered.

#### Barriers to Uptake of 99DOTS

##### Capability

Low literacy or limited language proficiency in available languages, more common among younger adolescents, impeded the ability to engage with 99DOTS and served as a barrier to uptake for some participants. In addition, healthcare providers further described that low literacy among adolescents increased the time to administer 99DOTS, as they had to provide more explanation and demonstration. This impacted the healthcare providers’ ability to fully explain or allow adolescents to engage and consider uptake of the approach.

> *“For those that knew how to use a phone, it was easy. Maybe for those who have not gone to school or do not know how to use a phone…it was hard because you know teaching someone how to use a phone when they have never used a phone before--”*-Healthcare provider, GH health facility

##### Opportunity

Limited agency in healthcare decision-making, variable and inconsistent mobile phone access, and connectivity were notable barriers to adolescents’ opportunity to engage in 99DOTS. A number of participants who did not engage with 99DOTS described how the approach was explained to their caregiver. Subsequently, the caregivers made the decision regarding enrollment regardless of the adolescents’ opinion, as described by an adolescent with TB who did not participate in 99DOTS (age 15-19):

> *“Yes, I wanted [to participate in 99DOTS] but because my mum was like ‘you will not keep up with it [99DOTS]’, I will not manage, I did not use it.”*

Although the quantitative analysis did not show any significant difference in uptake by sex, some healthcare providers noted that there was differential access to mobile phones among male and female adolescents, as described by a healthcare provider from an RRH facility: *“Poor access to the phone was more for females. Being adolescents, the parents could not allow the girls to use phones”.* Adolescent participants described limited or nuanced mobile phone access as a barrier to uptake. While older adolescents were more likely to have their own phone or more regular mobile phone access, the majority of adolescents described reliance on a family member or caregivers’ phone to use 99DOTS:

> *“My father was not always around. He promised to give me the phone whenever I needed to call. He became busy and he worked from far. My mother’s phone was with my grandmother. We did not go back to follow-up the process of making phone calls”* -Adolescent with TB, not enrolled on 99DOTS, 10-14 years old

##### Motivation

Disclosure of TB status was a concern among most adolescent participants regardless of the 99DOTS enrollment status. Adolescents who shared or used other people’s phones feared that 99DOTS calls or reminder messages could be sent when the phone was with another person to whom they were not ready to disclose their TB status. Some adolescents described this as a de-motivating factor in the uptake of 99DOTS.

> *“Maybe messages may be tricky because any one can see them. Even phones call for us who don’t have phones you may not know when the call will be made. I suggest that we agree with health workers about the time they can send a message or call”*-Adolescent with TB, did not participate in 99 DOTS, 15-19 years old

Some adolescents felt that they would be subjected to intensified monitoring by agreeing to use 99DOTS, with many expressing an underlying fear that healthcare providers would not be happy and would judge those whom the 99DOTS showed as non-adherent.

> *“They [healthcare providers] would judge you because they know that you are not taking your medicine and they can tell that this given patient is not.”* -Adolescent with TB, did not participate in 99 DOTS, 15-19 years old

Some healthcare providers described concerns about increased workload with the introduction of the 99DOTS platform.

> *Hmmm! Some of them [other healthcare providers] liked it, while others saw it as added work, and then others did not like it but did it because they had to do it. Others looked at it as added work but others liked it, as you know some people like it, but they do not like extra work as it that extra work involved.* -Healthcare provider, HC III health facility

## DISCUSSION

Digital adherence technologies such as 99DOTS can provide an option for remote TB treatment monitoring using a mobile phone and may be attractive to adolescents. As part of a 99DOTS scale-up across 30 facilities in Uganda, we found high uptake of 99DOTS among adolescents with TB (73%). Uptake was higher than for adults (52%) in the DOT for DAT trial (12). Uptake was higher among older adolescents and those with caregiver involvement and lower at higher-level facilities compared to HC IIIs. Qualitative findings further demonstrate that technological proficiency and literacy, access to training, caregiver involvement, and shared decision making influenced the uptake of 99DOTS. Limited mobile phone access, anticipated TB-related stigma and the workload of healthcare providers in hospitals posed barriers to 99DOTS uptake. Overall, adolescents are capable and motivated to participate in 99DOTS, but barriers linked to age, gender, social support, and health center/provider time and involvement present critical opportunity-related barriers that if addressed could further improve uptake and user experience.

Phone access and the ability to use a mobile phone played a role in the capability and opportunity for uptake of 99DOTS among participants in this study. The greater uptake among adolescents 15 to 19 years of age may be due to differences in phone accessibility, with more access among older adolescents compared to younger adolescents aged 10 to 14 years. While mobile phone access is widespread, ownership globally increases with age, with significantly higher phone ownership among older adolescents than younger (16); (17). Our qualitative findings also suggested disparate phone ownership and control by sex, with female adolescents potentially having lower phone ownership and control compared to male counterparts, which could impact participation in 99DOTS. The findings are consistent with a mixed methods study in Rakai (18), central Uganda, which found that complex gender disparities and control mechanisms may significantly influence adolescent and young women’s phone access.

Treatment supporters were essential in facilitating the opportunity for the uptake of 99DOTS among adolescents. The interviews revealed that treatment supporters provided additional explanations of 99DOTS to adolescents, were important facilitators of their ability to participate in 99DOTS by ensuring access to their mobile phones, and provided counseling on the use of 99DOTS. We found that caregivers can provide critical informational, instructional, and emotional support during TB treatment (19). These findings align with two studies from Peru among adolescents with TB. A qualitative study found that adolescent emotional support and treatment monitoring from caregivers were essential to adherence, and a prospective cohort study found that adolescents with the weakest social support had considerably worse adherence to treatment. Overall, studies have shown that adolescents benefit from the caregiver and parental support to facilitate decision-making and the ability to attend treatment follow-up schedules (3, 20, 21). Our findings add to this evidence by demonstrating the multifaceted pathways that treatment support can impact adolescent uptake of an intervention in the TB care setting.

Agency in health care decision-making limited adolescent uptake of 99DOTS, although adolescents reported motivation to participate in service of their recovery from TB. Adolescents 10 to 17 years are minors in Uganda and often dependent on parents or caregivers for decision support. Our qualitative findings show that these adolescents could have benefitted from the additional explanation about 99DOTS provided to caregivers. The role of caregiver counseling in adolescent decision-making is supported by other studies that focus on the parental role in improving uptake and adherence to TB treatment. A study among adolescents in Uganda suggested that parental sensitization by healthcare providers about TB improved involvement and care during treatment and is a key contributor to adolescent treatment uptake (22). However, our findings also highlight that while parents and caregivers are often facilitators, they may serve as barriers to intervention participation, as some participants in our study described not having the opportunity to decide whether to participate in 99DOTS. While different in context, the potential implications may not be dissimilar from the sexual and reproductive adolescent health decision-making literature, where adolescents may abstain from interventions or services because of the need for parental notification or involvement (23). Our findings demonstrate that an adolescent-centered approach that balances parental and adolescent decision-making and agency is needed to ensure adequate and sustainable enrollment and participation in digital adherence technologies. Both adolescents and healthcare providers raised concerns about inadvertent TB disclosure and stigma as motivational barriers to 99DOTS uptake. TB stigma, whether anticipated or experienced, is a significant concern for adolescents (24), and approaches to address or mitigate this concern must be taken into account as this will determine their uptake of 99DOTS regardless of agency.

This study had limitations. First, the study utilized secondary data collected in TB registers, which limited the number of variables evaluated. This was mitigated by conducting qualitative interviews to understand further 99DOTS beyond the variables recorded by the National TB register. Second, as the interviews were conducted after the completion of 99DOTS, only adolescents or their caregivers with working phone numbers or those whose location was known could be invited to participate in the study. However, study team members worked to obtain phone numbers for all individuals in the sampling frame who were selected for participation, including visiting health facilities, to obtain updated telephone information. Finally, these findings represent the experiences within the Kampala region of Uganda and thus may not be generalizable or transferrable to other settings. However, as Uganda and other countries adopt or scale up 99DOTS or similar digital adherence technologies, the themes from the qualitative findings can be helpful in planning and implementation.

## CONCLUSION

99DOTS-based treatment supervision had high overall uptake among adolescents treated for pulmonary TB, and healthcare providers were interested in and engaged in using the technology. 99DOTS is a promising digital platform for TB treatment adherence among adolescents, who often require tailored treatment approaches. 99DOTS implementation for adolescents can be enhanced with increased access to mobile phones and improved healthcare provider communication with adolescents and their caregivers to facilitate shared treatment decision-making and address TB-related stigma concerns. Policymakers and practitioners should leverage these facilitators and consider strategies to address barriers in the adolescent rollout of digital adherence technologies.

## METHODS AND MATERIALS

### Study Design

We used a sequential explanatory mixed methods design (25) to quantitatively identify factors associated with the uptake of 99DOTS among adolescents with DS-PTB using cross-sectional data (Phase 1) and further explore these factors in-depth using qualitative semi-structured interviews with adolescents diagnosed with DS-TB and health workers (Phase 2).

#### Theoretical framework

The Capability, Opportunity, Motivation, and Behavior (COM-B) model is a behavior change framework comprised of three constructs that interact to influence behavior (26). This model posits that to adopt a particular behavior (e.g., uptake of 99DOTS), there must be <u>c</u>apability, <u>o</u>pportunity, and <u>m</u>otivation to perform the behavior: behavior is defined as any action a person takes in response to internal or external events, capability is both physical and psychological ability or capacity to execute a behavior, opportunity is the external factors both physical and social that are associated with undertaking a specific behavior, and motivation is the automatic or reflective mental compulsion to carry out a specific behavioral option amidst alternatives (27).

#### Study Setting

The DOT to DAT trial and 99DOTS scale-up was conducted at 30 Designated TB treatment Units (DTU) including seven health center IIIs, six health center IVs, and 17 hospitals in the Kampala, Uganda region. These health facilities are located in urban Kampala metropolitan and rural regions with a radius of 280 kilometers outside Kampala. Health center IIIs are located at the sub-county level and offer basic emergency care, while health center IVs are larger and located at health sub-districts and provide comprehensive care in addition to other primary health care services offered at Health center III. Hospitals include both general and regional referral hospitals. The complexity of cases handled increases from Health Center III to regional referral hospitals. Health facilities were selected based on their TB notification rates in the Kampala region (MOH, 2020).

#### Study Population

Adolescents 10-19 years of age with DS-PTB treated with adult fixed-dose combination (2 RHZE; 4RH) at the 30 DOT to DAT trial and 99DOTS scale-up TB treatment sites in Uganda from July 2020-June 2021 were eligible for inclusion in quantitative analyses. All adolescents from the quantitative sample and all health workers were eligible for qualitative semi-structured in-depth interviews.

#### Sample size

The sample size was fixed (N=410) based on the number of adolescents treated for TB at participating health centers during the 99DOTS scale-up project period.Drawing upon maximum variation sampling on the primary descriptors of interest from the quantitative data (age, sex, HIV status, and level of care), a sample size of 20 adolescents was targeted for qualitative in-depth interviews and eight health workers for key informant interviews (28). The study anticipated saturation would be achieved, given that respondents were relatively homogenous within their sampling categories (28). During qualitative data collection, study data was monitored through weekly team calls and debriefing to assess saturation.

### Phase 1 (Quantitative)

#### Sampling

Sampling for the DOT to DAT trial is described elsewhere (12). All adolescents 10 to 19 years who were initiated on anti-TB treatment during the scale-up phase of the DOT to DAT study were eligible for inclusion in the quantitative analysis. Adolescents who had incomplete baseline treatment records were excluded because the study could not ascertain the treatment supervision approach used. The study also excluded those diagnosed with TB osteomyelitis, meningitis, or multi-drug resistant TB because they were not eligible for enrollment on 99DOTS.

#### Data collection

Routine patient data from TB treatment centers entered into the National TB register for all study sites was abstracted by study staff into a REDCap database (29). The DOT to DAT trial collected additional data on patient age, weight, height, address, telephone number, diagnostic and treatment facility, clinical status, HIV status, treatment issued, and TB treatment outcomes.

#### Study Variables

##### Independent variables

Independent variables included adolescent social and demographic characteristics and health system factors that may influence the uptake of 99DOTS: sex, level of health care, presence of treatment supporter at diagnosis, HIV status, and type of TB disease being managed. Sex, HIV serostatus, and category of TB disease were classified as binary categorical variables. Age in years was categorized as either “younger adolescents” (10 to 14 years) or “older adolescents” (15 to 19 years), and level of health care was classified as a categorical, ordinal variable with health centers (levels III, IV), hospitals, and regional referral hospitals as different levels of care in-line with determinations by the Ministry of Health in Uganda(30).

##### Dependent variables

The primary outcome variable was the uptake of 99DOTS. We defined the primary outcome as binary: received 99DOTS or routine CB-DOT.

#### Statistical Analyses

We used descriptive statistics to summarize participant characteristics. We used a modified poison regression (31) to determine the association between age, sex, level of health care, HIV serostatus, type of TB disease, and availability of a treatment supporter, with uptake of 99DOTS. The final multivariate model was fitted with variables based on the extant literature (age, sex) or factors associated with uptake of 99DOTS at *p*<0.20 in univariate analysis (32). Further, collinearity within the predictor variables was ruled out, and a stepwise approach was used to build the multivariable model and model fitness evaluated (Pearson’s chi-squared p-value=0.87). Data were analyzed using Stata version 14.3 (StataCorp, College Station) (33).

### Phase 2 (Qualitative)

#### Sampling

Using a sampling frame of adolescent patients treated for DS-PTB at participating 99DOTS scale-up TB treatment centers, we aimed to recruit approximately even numbers of adolescents who participated in 99DOTS and adolescents who declined to participate in 99DOTS. In addition, the health workers who managed participating TB treatment centers and provided care to adolescents for TB during the study period were purposively approached for participation in key informant interviews.

#### Data collection

In-depth interviews (IDIs) were conducted by experienced Ugandan interviewers using semi-structured guides. Interviewers received additional training on the study protocol and adolescent engagement. Interview guides were developed to elicit experiences with 99DOTS, including barriers and facilitators to enrollment and its use. Questions were informed by DOT to DAT trial findings and the COM-B model (27) and explored perceptions of 99DOTS, understanding of 99DOTs, and factors that might influence 99DOTS uptake. In addition to these questions, health workers were asked for their perceptions of the training they received and their use of 99DOTS. IDIs with adolescents and health workers were conducted by trained qualitative interviewers in the participant’s language of choice (English, Luganda, or Lusoga). Interviews were held at health facilities or by telephone, per participant preference. Interviews lasted between 30 and 60 minutes, were audio-recorded, transcribed verbatim, translated into English (as necessary), and uploaded into Dedoose version 4.3 for data management, coding, and analysis (34).

#### Analysis

We utilized a thematic analysis approach to explore pre-defined areas of interest related to 99DOTS uptake from the quantitative findings and prior theoretical frameworks while allowing detailed and new insights and themes to emerge from the data (35). The study analysis team developed an initial codebook consisting of a-priori codes informed by quantitative findings and the COM-B domains. Study team members read all transcripts and open-coded a subset of transcripts in parallel to further identify codes. The codebook was iteratively refined during the initial data familiarization and coding process, with inductive codes added to reflect emergent themes. When the codebook was finalized, all codes were systematically applied across all the transcripts, starting with double coding completed on a subset of four transcripts until consensus on code domains and definitions was reached among analysis team members. Following coding, data were organized into a chart format for each key theme that included summaries of different barriers and facilitators of uptake of 99DOTS among adolescents with respective illustrative quotes. Themes were iteratively compared and explored across participant types and sub-categories (age, sex, HIV status) to facilitate the identification of potential explanations for findings from the quantitative analysis. The themes were further situated within the COM-B model (26, 27) to provide a framework to inform understanding of uptake of 99DOTS in the context of barriers and facilitators.

## Data Availability

Data, that supports the findings of this study are available from the corresponding author. It is available on open science frame work platform. https://osf.io/k2mq6/files/osfstorage/67498957a08e49e00d6facea

https://osf.io/k2mq6/

https://osf.io/k2mq6/files/osfstorage/67498957a08e49e00d6facea

## Author Contributions

Conceptualization, A.C, A.K, D.J, A.K; Methodology, A.C, A.K, D.J; Validation A.C, A.K, D.J; Investigation, P.W, N.W, J.N, A.K, R.C, L.K; Formal Analysis P.W, N.W, J.N, A.K; Writing-Original Draft Preparation, P.W, N.W, J.N ; Writing-Review & Editing, P.W, N.W, J.N, E.W, A. A.C, A.K, D.J; Supervision, A.C, A.K, D.J, N.W, J.N, E.W ; Project Administration, A.C, A.K, D.J, N.W, J.N, E.W; Funding Acquisition, A.C, A.K.

## Funding

This study was supported by the Stop TB Partnership’s TB REACH initiative, grant number STBP/TBREACH/GSA/W6-37 (AC, AK), which is funded by the Government of Canada, the Bill & Melinda Gates Foundation, the United States Agency for International Development and Makerere University College of Health Sciences -University of carlifornia Berkeley-Yale Pulmonary Complications of AIDS Research Training (PART) Program, NIH D43TW009607, from the Forgaty International Center (FIC).

## Conflict of interest

The Authors declare no conflict of interest.

## Ethics Standards Disclosure

The study obtained approval from the Makerere University School of Public Health Research and Ethics Committee to conduct the sub-study (Protocol #630) and Uganda National Council for Science and Technology (HS 2436). Further, informed consent was obtained from all participants for in-depth and key informant interviews and assent for adolescents under 18 years old.

## Notes

### Competing Interest Statement

The authors have declared no competing interest.

### Author Declarations

1) Makerere University School of Public Health Research and Ethics Committee -Protocol #630 2) Uganda National Council for Science and Technology -HS 2436.

